# Motor and cognitive outcomes of low birth weight neonates born in a limited resource country: a systematic review

**DOI:** 10.1101/2020.08.31.20178277

**Authors:** Graciane Radaelli, Eduardo Leal-Conceição, Felipe K. Neto, Melissa R. G. Taurisano, Fernanda Majolo, Fernanda T. K. Bruzzo, Booij Linda, Magda L. Nunes

## Abstract

**Objective:** To examine the outcomes of motor and cognitive development among neonates born with low birth weight (LBW) or very low birth weight (VLBW).

**Data sources:** Systematic review carried out in PubMed, Cochrane Library and Web of Science using the search strategy using combinations of the following keywords and terms: preterm birth OR prematurity OR premature Infants OR premature children AND low birth weight children OR very low birth weight children AND neurodevelopment OR cognitive development OR Motor development OR follow up AND humans. Articles searched were published from inception until July, 2019, and involved children born and evaluated in Brazil. The bias risk analysis was adapted from the STROBE scale, used to evaluate the methodology of the included studies.

**Data synthesis:** The search identified 2,214 publications. After screening for titles and abstracts and removing duplicate entries, full texts of 38 articles were reviewed. After reading full texts, 24 articles met the inclusion criteria (articles in Portuguese and English), dated from 1998 to 2017). Endnote Version X9 software was used for data extraction. Two reviewers performed the literature search and study selection independently. Disagreements were solved by consensus or by a third reviewer.

**Results:** it was evidenced an inferior motor development of children with LBW when compared to the control population, the standardized mean difference of [-1.15 (95% CI - 1.56, -0.73), I^2^ 80%], children with LBW have lower cognitive development according the standardized mean difference of [-0,71 (95% CI -0.99, -0.44) I^2^ 67%].

**Conclusion:** Our review reinforces that impaired motor and cognitive outcome is a significant long-term outcome associated with LBW. The risk of impairment in those domains increases with decreasing gestational age.

## Introduction

Strong evidence shows that children born prematurely or small for gestational age have a greater predisposition to deficits and / or delayed neuropsychomotor development^1^ and deficit rates are inversely related to gestational age and birth weight.^2^

Prematurity is a growing health problem worldwide, especially in developing countries where access to obstetric service and neonatal support is not guaranteed to the entire population.^3^ Worldwide 965.000 deaths occur in the neonatal period and 125.000 deaths between 1-5 years of age due to prematurity, representing the leading cause of neonatal and infant death.^4^ The worldwide incidence of deliveries before 37 weeks of gestation is 11.1% with large geographical differences, ranging from 5% in developed countries to 18% in countries with less economic power.^5^ In South America, the mortality of children born with very low birth weight reaches 26%,^5^ demonstrating their socioeconomic lability.

Intrauterine growth restriction (IUGR) is an abnormal fetal growth standard that happens in approximately ten percent of gestations and is related with neonatal morbidity and mortality^6^ IUGR is a condition in which the fetus does not reach the expected weight during pregnancy, is defined as a rate of fetal growth that is less than normal for the expected growth potential of a particular infant, because of genetic or environmental factors,^7^ it is a pathologic inhibition of intrauterine fetal growth and the failure of the fetus to achieve its growth potential^8^. In IUGR pregnancies, the fetus attempts to prevent damage by slowing its growth and shortening its gestation, causing prematurity.^6^

The terms IUGR and small for gestational age (SGA) have been used as equals in the literature, although there is a difference. IUGR reflects fetal suffering, though SGA only provides a measure of size and not a measure of antenatal growth quality.^6^ SGA is a term used to describe neonates whose birth weight is bellow than the usual amount for the number of weeks of pregnancy, they usually are below the 10th percentile for babies of the same gestational age, or 2 deviations below.^7^

Subjects born with IUGR experience several poorer developmental outcomes, involving cognitive, socioemotional and behavioral domains, compared with children born appropriate for gestational age.^6^ IUGR is one of the main causes of perinatal and neonatal morbidity and mortality and contributes to long-term chronic diseases.^7-9^ Neurodevelopmental impairment, immediate perinatal adverse events (prematurity, cerebral palsy, intrauterine fetal death, neonatal death) and also to adult pathologic conditions are leading consequences of the IUGR.^8,10^ These neonates have different types of short-term and long-term problems making them vulnerable, both need immediately and long-term follow-up, especially for delayed onset of neurological disorder so that a quick intervention can be initiated on neurological and physical aspect and proceed to better disclosure.^9^

The exposure to undernourishment early in life appears to have detrimental effects on the developing brain.^11-13^ Experimental studies conducted on animal models of early malnutrition and epilepsy, showed that malnutrition per se has effects not only on hippocampal morphology such as decreased neuronal density in CA1 and CA3 subfields and decreased NDMA expression during adulthood^11,12^ but also leads to progressive learning and memory disability.^1^ However, the enriched environment led to a significant benefit in learning and retention of visual-spatial memory being able to reverse the cognitive impairment.^13^

The average intelligence quotient (IQ) of a child with low birth weight (LBW) is lower than a child with adequate birth weight.^14^ In addition to the reduced IQ, there are also greater learning difficulties and a greater need for special education in small for gestational age (SGAs) and preterm infants than in the general population.^15^ Cognitive deficits, school difficulties, and behavioral problems are often reported by children born with LBW or extreme preterm infants.^1^ While deficits have been reported across many cognitive domains, attention deficit is an area of concern for parents and teachers of premature children.^16^ Research shows that children born prematurely are much more at risk for neurobehavioral deficits even though there are individual variations and varying profiles and severity of deficits.^1^

The data concerning the medium or long-term outcomes of preterm newborns in South American countries remain deficient, and specific follow-up programs are challenging to provide in public institutions. After discharge from a neonatal intensive care unit (NICU), most of these infants are followed at primary care facilities, leaving most of them without specific attention.^17^

Given the great impact of low birth weight and/or prematurity on aspects of children neurodevelopment, it is extremely necessary to carry out studies that investigate these phenomena in greater depth in more heterogeneous populations like the ones from limited resources countries. The aim of this study was to examine the cognitive and motor outcomes of children with LBW or very low birth weight (VLBW), from studies conducted with the Brazilian population. Our hypothesis is that being raised in a limited resource country might have additional negative effects in the outcomes studied.

## Methods

This systematic review followed the criteria of the Preferred Reporting Items for Systematic Reviews and Meta-Analyses Checklist (PRISMA).^18^ The protocol of this systematic review was registered in international prospective register of systematic reviews (PROSPERO) under number CRD42019112403.

### Eligibility criteria

The criteria used for study inclusion were: original articles that established an association between gestational weight (GA), birth weight (BW), and neurodevelopment in Brazilian children; studies published from inception until July 2019, in Portuguese and English, and that used the cohort, case-control, longitudinal, cross-sectional, descriptive analytical, and retrospective methods. The dependent variables used in this review were the variables obtained as the result of tests (receptive language and/or expressive language). The independent variables were GA, BW, gender, age at time of evaluation, family income, and maternal level of education. For the purposes of the present study, prematurity was considered at three levels: borderline preterm (GA 35-36 weeks), moderately preterm (GA 31-34 weeks), and extremely preterm (GA ≤ 30 weeks). Newborns with low BW were classified as LBW (< 2,500 g), VLBW (< 1,500 g), and ELBW (< 1,000 g).

### Research strategies

A systematic review was carried out in the PubMed, LILACS and SciELO databases, using combinations of the following keywords and terms: preterm birth OR prematurity OR premature Infants OR premature children AND low birth weight children OR very low birth weight children AND neurodevelopment OR cognitive development OR Motor development OR follow up AND humans.

### Data Synthesis

Endnote Version X9 software was used for data extraction. The databases were searched and duplicate entries were removed. Abstracts that did not provide sufficient information regarding the inclusion and exclusion criteria were selected for full-text evaluation. In the second phase, the same reviewers independently evaluated the full text of these articles and made their selection in accordance with the eligibility criteria. Two reviewers (M.R.T. and F.T.B.) performed the literature search and study selection independently. Disagreements were solved by consensus or by a third reviewer.

### Risk of bias in individual studies

Two authors (M.R.T. and F.T.B.) were in charge of reviewing the methodological quality and the risks of bias according to the scale adapted from STROBE Statement^19^, considering only the studies that fit the inclusion criteria. A third author (G.R.) evaluated and defined any disagreements. The STROBE Statement scale aims to evaluate studies not related to randomized clinical trials; it comprises 22 applicable questions/items to assess the quality and biases of articles. These criteria assess the quality of data, internal validity (biases and confounding factors), external validity, and the ability of the study to detect a significant effect. To assess the risk of bias using the STROBE criteria, the articles of this systematic review were grouped into three different categories, each with a specific score: (a) first category: articles involving prevalence-type cross-sectional studies, with a maximum score of 12; (b) second category: articles with a cross-sectional and cohort methodological design, with a maximum score of 22; (c) third category: articles involving case-control studies, with intervention and maximum score of 22. To guarantee the proportion of results between the categories, the score obtained from each article was divided by the maximum possible score for each of the three established categories.

### Statistical analysis

We performed random effects calculation of weighted estimated average Odds Ratios (OR). Heterogeneity and publication bias were assessed with the I^2^ metric and funnel plots/Egger’s test, respectively. For continuous outcomes, if the unit of measurement was consistent throughout trials, results were presented as weighted mean difference with 95% of confidence intervals (CIs). Calculations were performed using random effects method and the statistical method used was inverse variance. Statistical significance defined for the analyzes as p < 0.05. Statistical heterogeneity of the treatment effects among studies was assessed using Cochran’s Q test and the inconsistency I^2^ test. Statistical heterogeneity of the treatment effects among studies will be assessed using the Cochran’s Q test and the inconsistency I-squared test, in which values above 25% and 50% are considered to be indicative of moderate and high heterogeneity, respectively.^20^

## Results

### Study selection

The initial database search resulted in 2409 articles. After removing the duplicate files, 2279 articles were filtered according to our inclusion criteria, from that 2217 were excluded by analyzing the titles and abstracts, resulting in 62 articles for evaluation of the full text. From the articles remained, 25 articles were selected and included in this review.^21-44^ The flowchart is shown in Fig. 1.

**Fig. 1.**
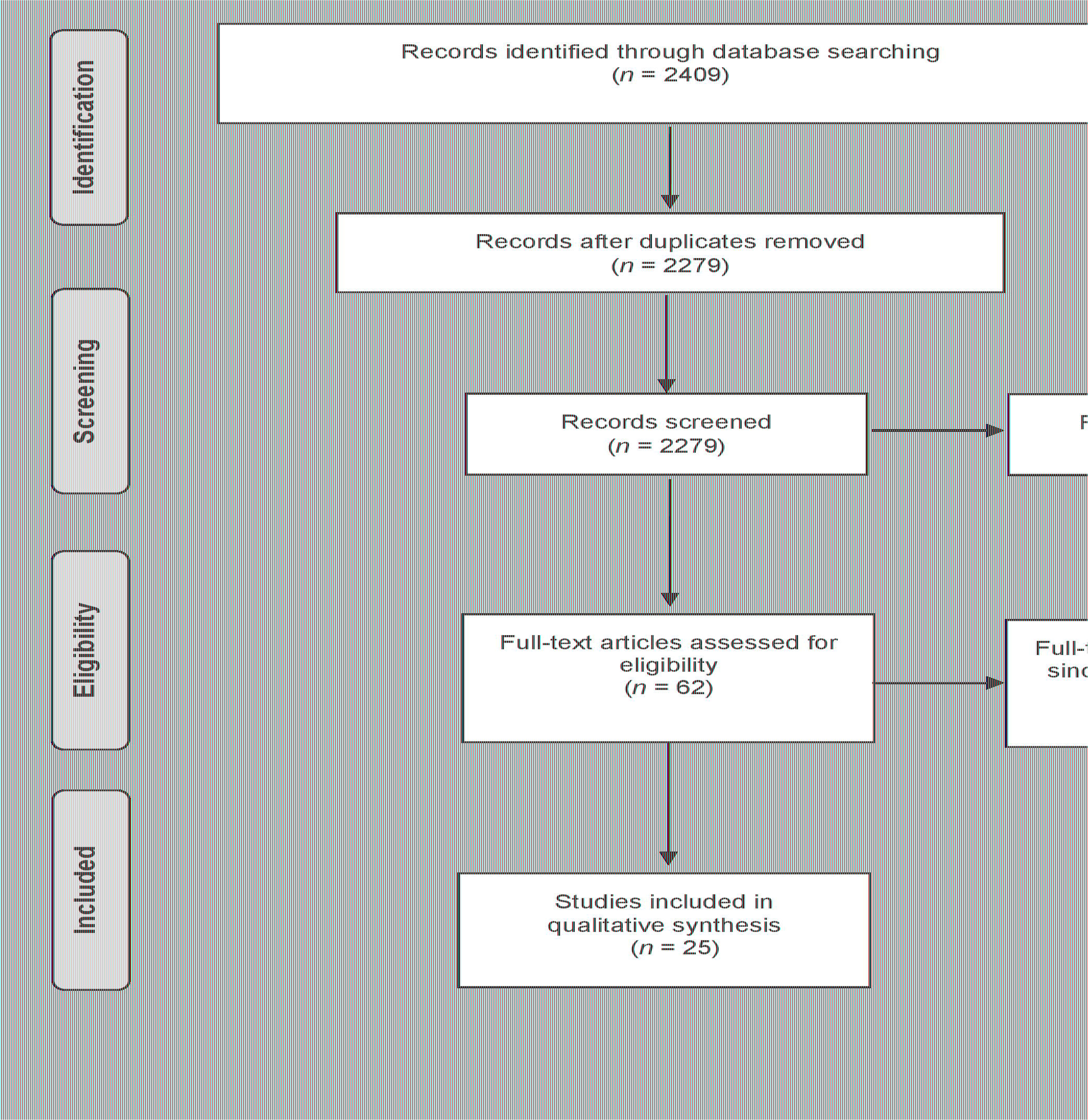
Summary of evidence search and study selection.

### Study characteristics

Included studies were published between the years 1998-2017. The average sample size in the aforementioned studies was 95.64 (SD=48.95), ranging from 32-262 participants. Approximately 52,8% (SD=7,8%) of the included samples were females. The mean birth weight of the premature participants was 1.477 kg, with standard deviation ± 0.424 and range 1.058-2.346 kg. The mean gestational age in the premature children was 32±4.2 weeks (range: 23-33.2 weeks). Motor and cognitive assessments were performed at 29.2 ± 24.5 (0-96) months of birth. The details of the individuals studies are shown in Table 1.

**Table 1.** Characteristics of the studies included in this systematic review.

| Author | Year | Birth weight | Gestational age<br>in weeks | Age of assessment<br>(months) | Proportion<br>girls | N |
| --- | --- | --- | --- | --- | --- | --- |
| Mello et al. | 1998 | 1185 | 32.2 | 21 | 0.65 | 70 |
| Grantham-McGregor et al. | 1998 | 2338 x 3210 | 37 x 37 | 6 to 12 | 0.615 | 262 |
| Mello et al. | 1999 | 1176 | 32 | 12 to 30 | - | 83 |
| Eickman et al. | 2002 | 3255 x 2332 | $\geq 37$ | 24 | 0.58 | 152 |
| Meio et al. | 2003 | 1903 | 32.04 | 48 to 60 | 0.64 | 79 |
| Méio et al. | 2004 | 1220 | 32 | 0 to 36 | 0.62 | 129 |
| Emond et al. | 2006 | 2346 x 3212 | 39 x 40 | 96 | - | 164 |
| Schirmer et al. | 2006 | 1622 x 1483 | 33.18 x 32.03 | 36 | 0.53 | 69 |
| Carvalho et al. | 2008 | 1058 | 30.44 | 12 | 0.5 | 36 |
| Manacero et al. | 2008 | 1417 x 2090 | 32.4 x 33.2 | 4 to 8 | 0.34 | 88 |
| Espirito Santo et al. | 2009 | 1788 | 32.3 | 48 to 60 | 0.5 | 80 |
| Mello et al. | 2009 | 1126 | 29.6 | 12 | 0.53 | 100 |
| Bühler et al. | 2009 | 3291 x 1073 | 39.1 x 29.4 | 24 | 0.43 | 32 |
| Magalhães et al. | 2009 | 3215 x 1171 | 39.97 x 30.23 | 84 | 0.65 | 70 |
| Procianoy et al. | 2009 | 1130 x 1250 | 31.7 x 29.3 | 24 | 0.49 | 131 |
| Silva et al. | 2011 | 1236 | 31 | 24 | 0.42 | 69 |
| Oliveira et al. | 2011 | 3273 x 1201 | 39 x 30 | 60 to 72 | 0.6 | 46 |
| Guimarães et al. | 2011 | 3158 x 1424 | 38-40 x 23-33 | 36 | 0.5 | 92 |
| Reis et al. | 2012 | 1122 | 29 | 24 | 0.53 | 109 |
| Fernandes et al. | 2012 | 1172 | 30 | 18 to 2 | 0.51 | 58 |
| Lemos et al. | 2012 | 1439 | 31 | 2 to 7 | 0.51 | 98 |
| Fan et al. | 2013 | 1890 | 33,6 | 6 to 7 | 0.5 | 97 |
| Guerra et al. | 2014 | 1727 x 1762 | 33.3 x 33.2 | 18 to 24 | 0.51 | 100 |
| Fuentefria et al. | 2017 | 1098 x 3348 | 29.1 x 39.1 | 8 to 18 | 0.48 | 135 |
| Saccani et al. | 2017 | 1886 X 2878 | 36 x 36 | 0 to 12 | - | 42 |

### Overview of functional relations with cognitive development in preterm

Of the included studies, 12 (48%) were cohort studies, (12) 48% cross sectional studies and 1 (4%) was a case-control study. Among cohort studies, 6 studies (50%) had a control group and 6 studies (50%) did not have a control group. In the cross-sectional studies, only 4 studies (33.3%) used a control group and 8 studies (66.7%) did not. The only case-control study that was included used a control group (100%). Most studies (19/76%) evaluated cognitive and motor outcomes, 4 studies (16%) only verified motor outcomes and 2 studies (8%) only cognitive outcomes.

The most commonly used instruments used were: Bayley Scales of Infant Development, present in 13 (54.1%) studies; Home Observation for Measurement of the Environment and Denver Developmental Screening Test, both present in four studies (16.6%). An overview of functional relations with cognitive development in preterm/ low birth weight /very low birth weight children about the studies were demonstrated in Table 2.

**Table 2.**
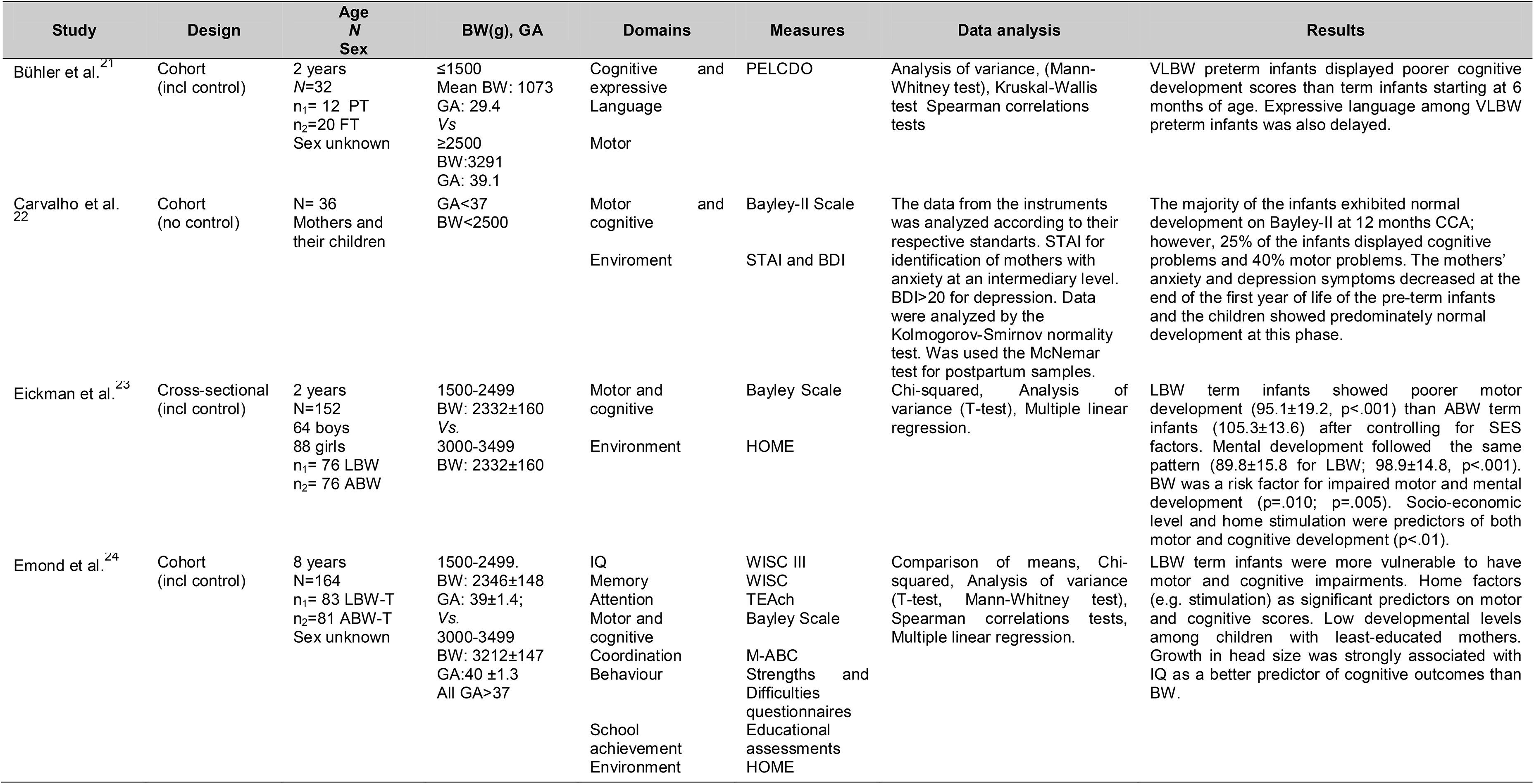
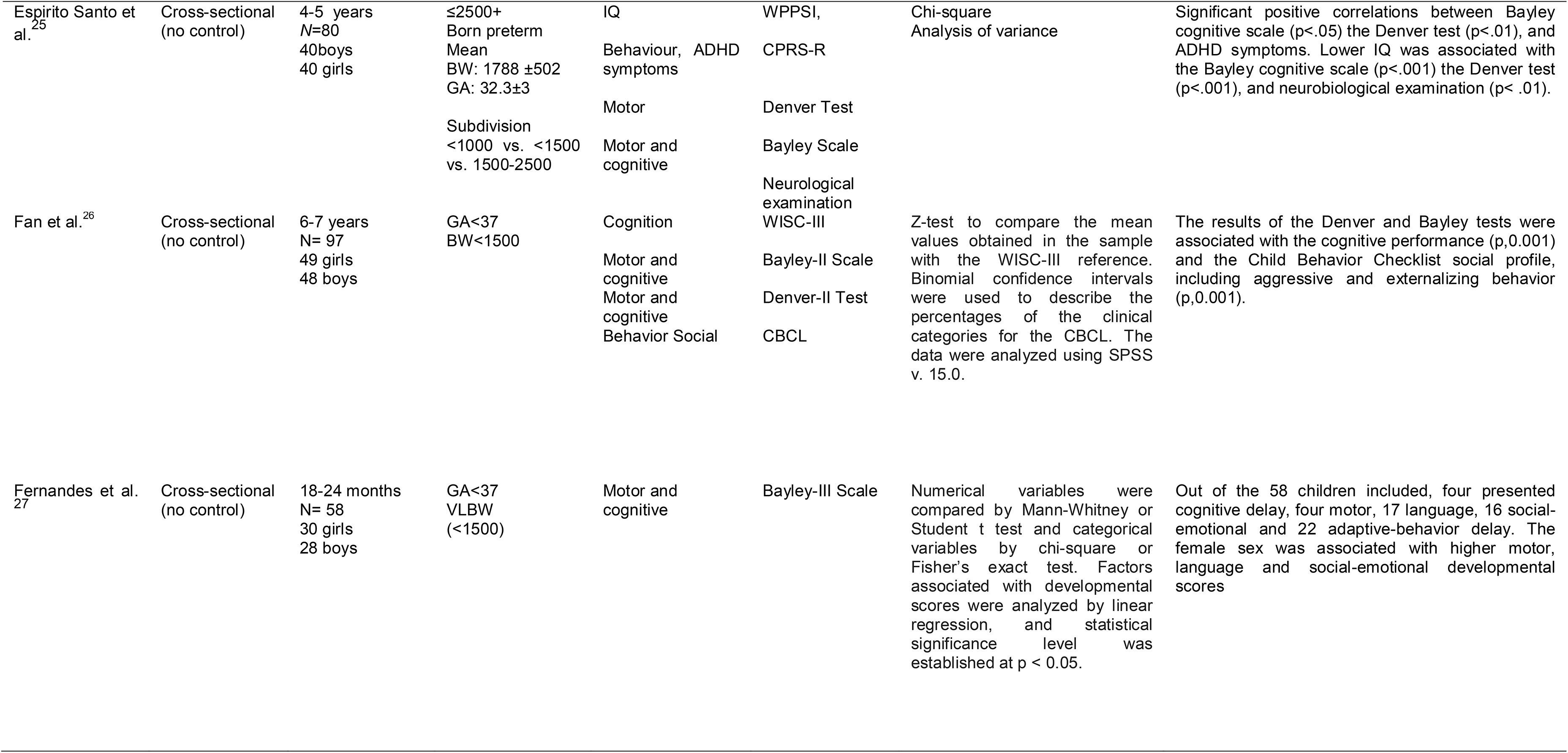
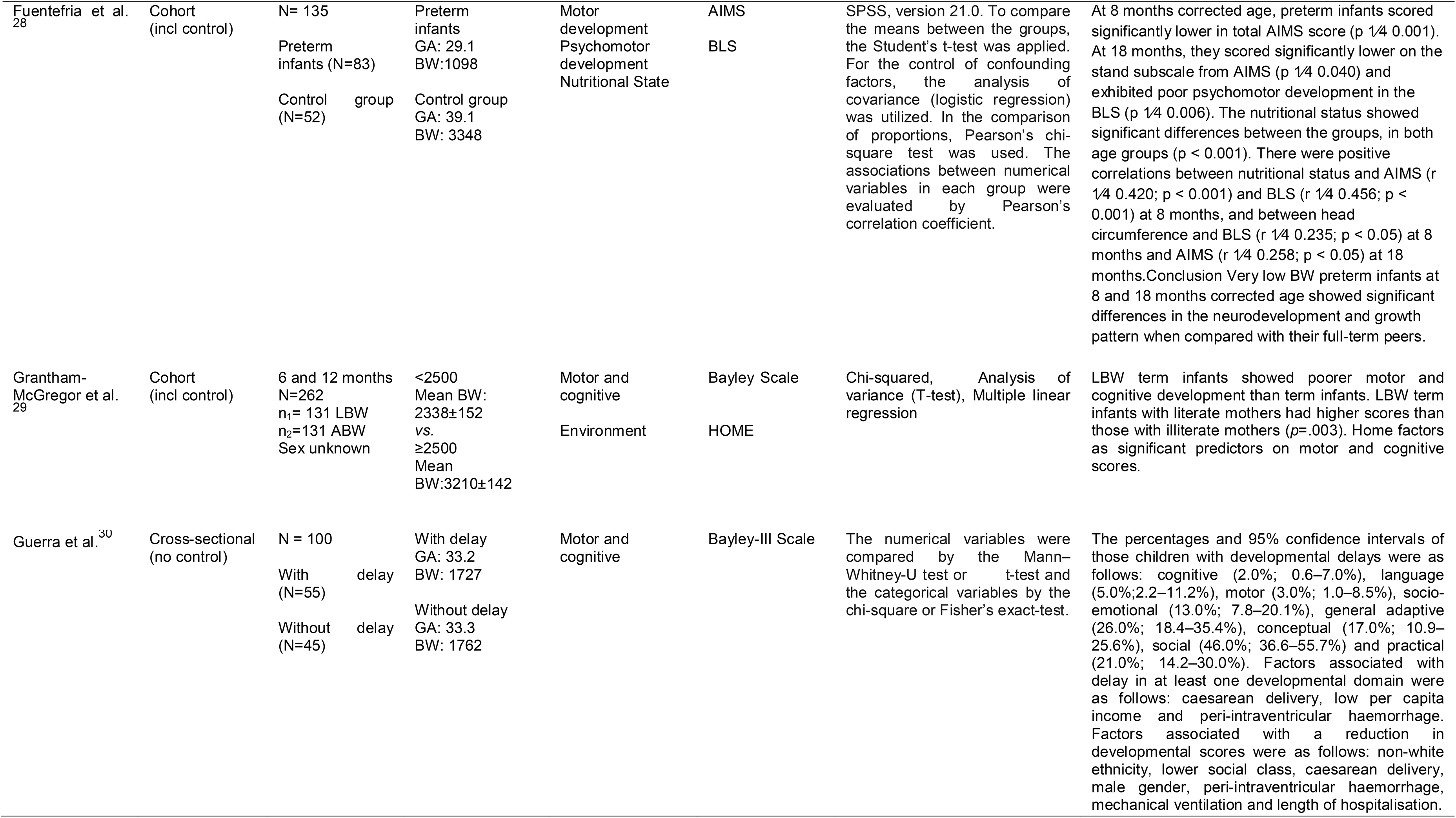
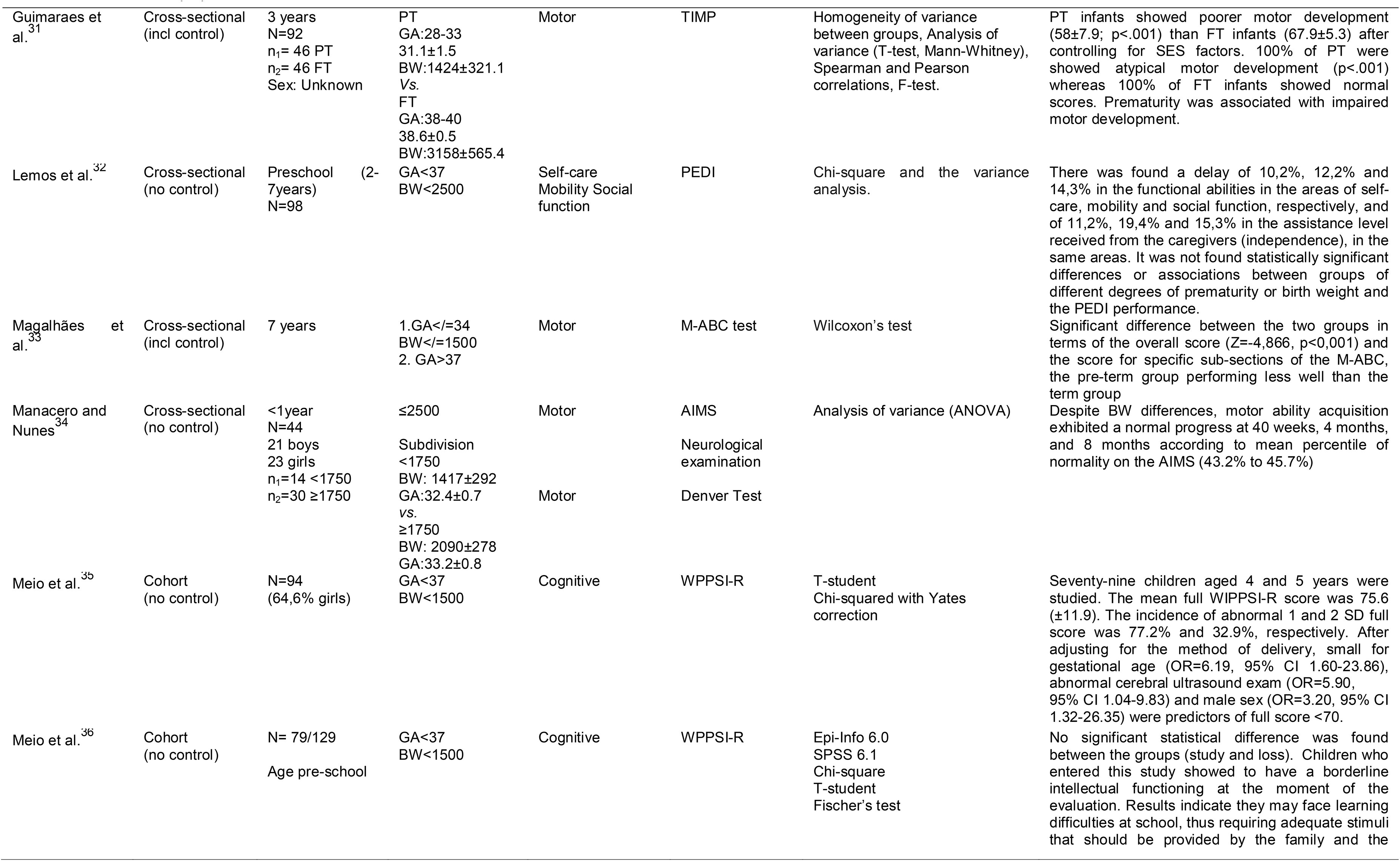
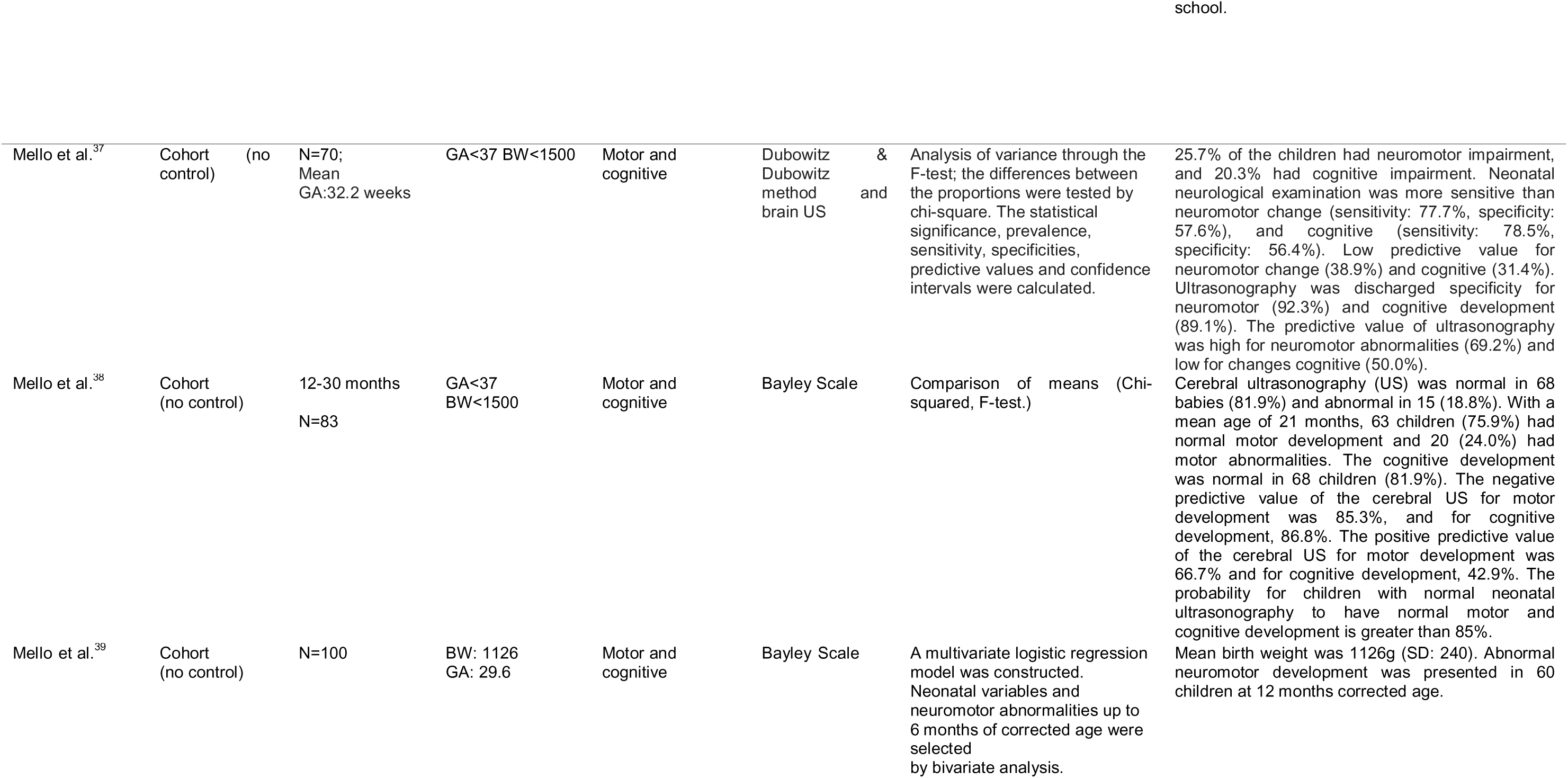
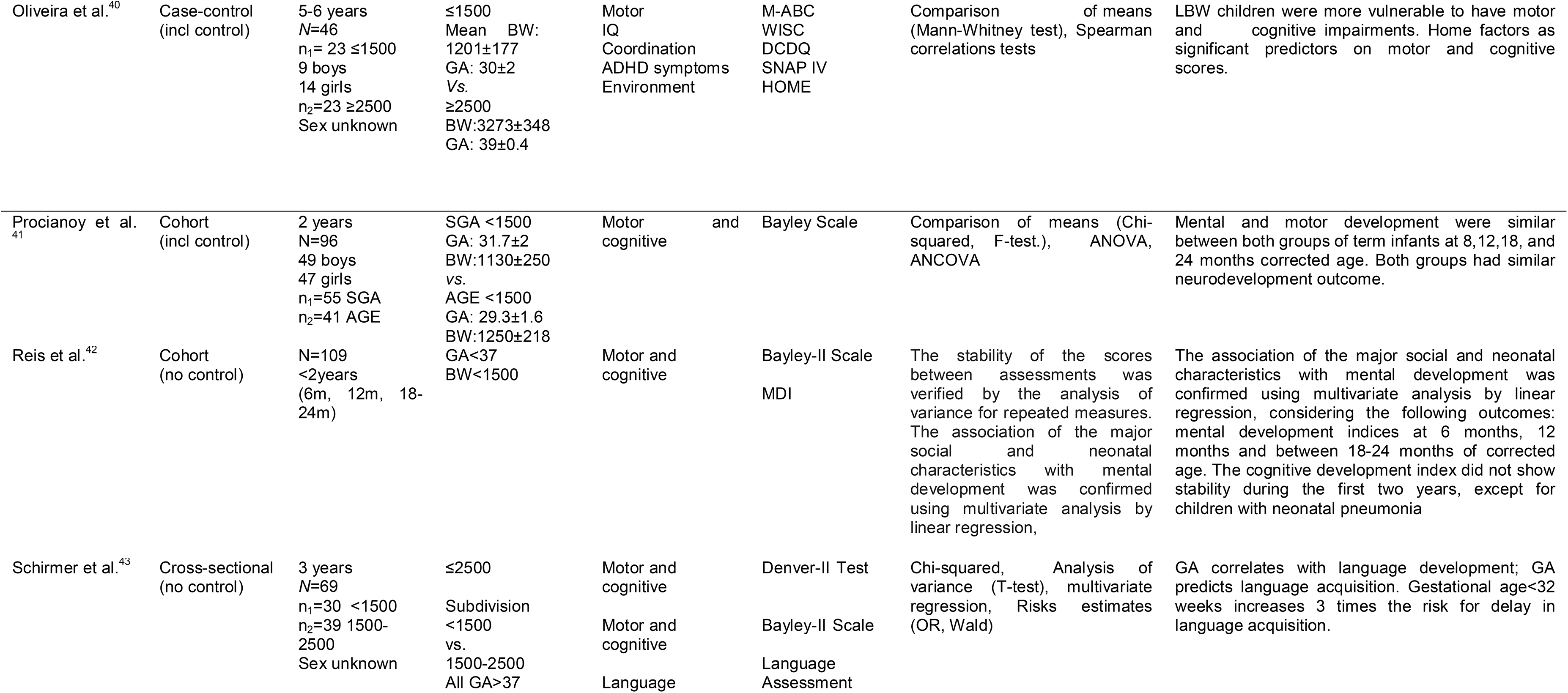
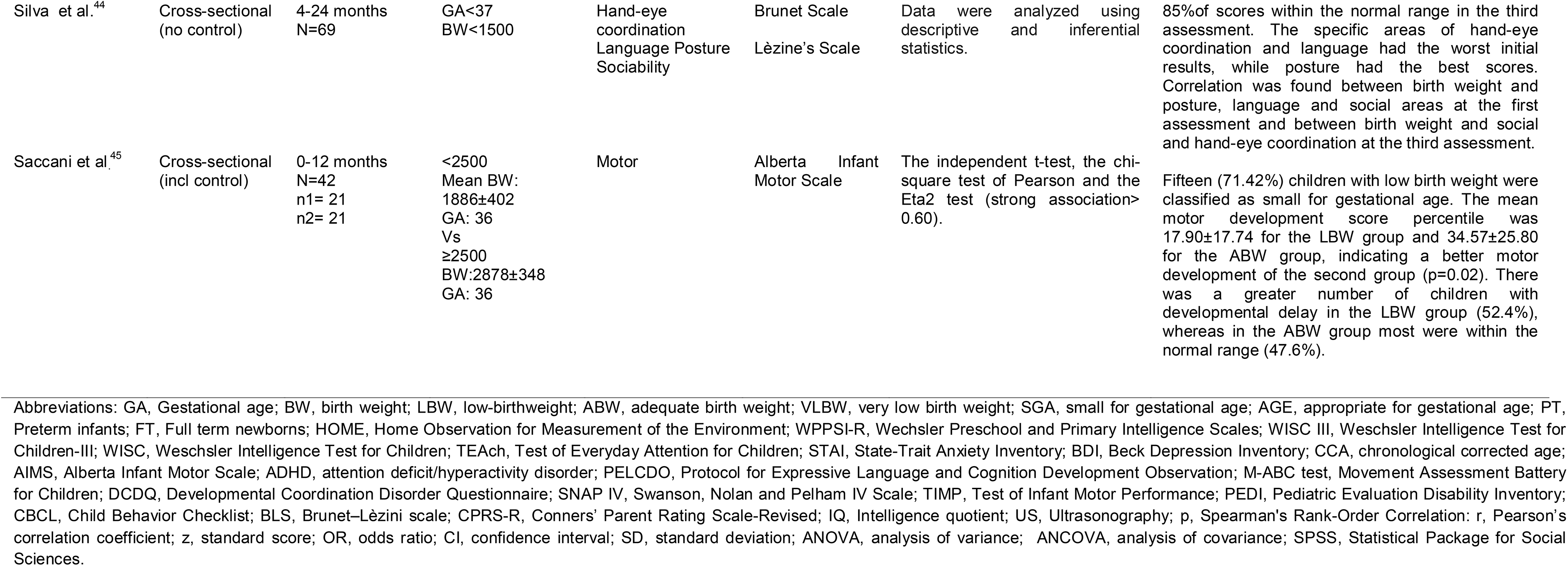
Overview of functional relations with cognitive development in preterm/ low birth weight /very low birth weight children.

### Risk of bias assessment about studies shows

The assessment of the methodological quality and risk of bias is shown in Table 3. Of the 25 articles evaluated, a mean score of 93.16 (±3.9)was obtained, with a maximum score of 100.0% and a minimum score of 86%. Thirteen articles showed values below the mean score and, therefore, were considered as lower methodological quality.

**Table 3.**
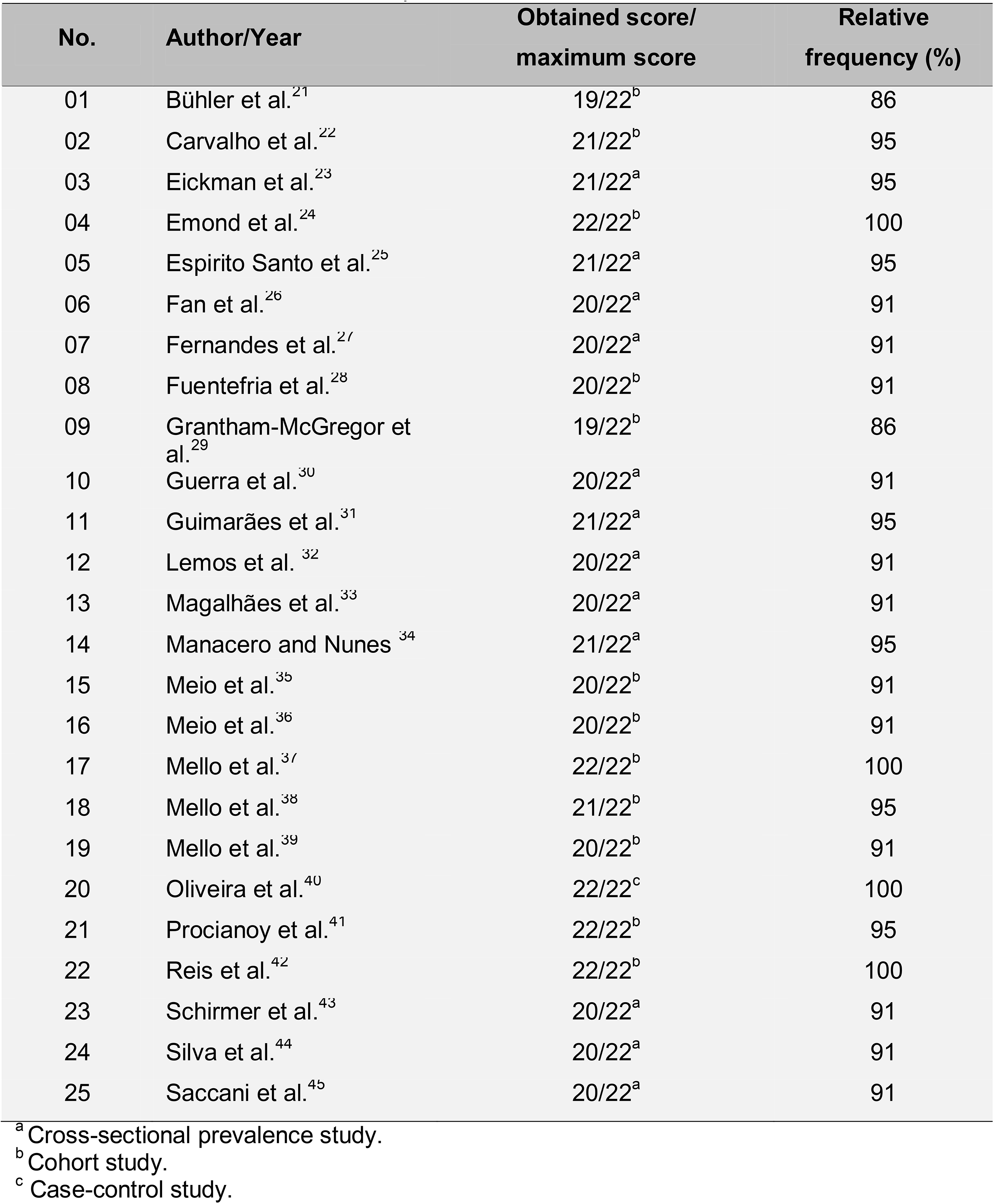
Risk of bias assessment adapted from STROBE.^19^

| No. | Author/Year | Obtained score/<br>maximum score | Relative<br>frequency (%) |
| --- | --- | --- | --- |
| 01 | Bühler et al.<sup>21</sup> | 19/22<sup>b</sup> | 86 |
| 02 | Carvalho et al.<sup>22</sup> | 21/22<sup>b</sup> | 95 |
| 03 | Eickman et al.<sup>23</sup> | 21/22<sup>a</sup> | 95 |
| 04 | Emond et al.<sup>24</sup> | 22/22<sup>b</sup> | 100 |
| 05 | Espirito Santo et al.<sup>25</sup> | 21/22<sup>a</sup> | 95 |
| 06 | Fan et al.<sup>26</sup> | 20/22<sup>a</sup> | 91 |
| 07 | Fernandes et al.<sup>27</sup> | 20/22<sup>a</sup> | 91 |
| 08 | Fuentefria et al.<sup>28</sup> | 20/22<sup>b</sup> | 91 |
| 09 | Grantham-McGregor et al.<sup>29</sup> | 19/22<sup>b</sup> | 86 |
| 10 | Guerra et al.<sup>30</sup> | 20/22<sup>a</sup> | 91 |
| 11 | Guimarães et al.<sup>31</sup> | 21/22<sup>a</sup> | 95 |
| 12 | Lemos et al.<sup>32</sup> | 20/22<sup>a</sup> | 91 |
| 13 | Magalhães et al.<sup>33</sup> | 20/22<sup>a</sup> | 91 |
| 14 | Manacero and Nunes<sup>34</sup> | 21/22<sup>a</sup> | 95 |
| 15 | Meio et al.<sup>35</sup> | 20/22<sup>b</sup> | 91 |
| 16 | Meio et al.<sup>36</sup> | 20/22<sup>b</sup> | 91 |
| 17 | Mello et al.<sup>37</sup> | 22/22<sup>b</sup> | 100 |
| 18 | Mello et al.<sup>38</sup> | 21/22<sup>b</sup> | 95 |
| 19 | Mello et al.<sup>39</sup> | 20/22<sup>b</sup> | 91 |
| 20 | Oliveira et al.<sup>40</sup> | 22/22<sup>c</sup> | 100 |
| 21 | Procianoy et al.<sup>41</sup> | 22/22<sup>b</sup> | 95 |
| 22 | Reis et al.<sup>42</sup> | 22/22<sup>b</sup> | 100 |
| 23 | Schirmer et al.<sup>43</sup> | 20/22<sup>a</sup> | 91 |
| 24 | Silva et al.<sup>44</sup> | 20/22<sup>a</sup> | 91 |
| 25 | Saccani et al.<sup>45</sup> | 20/22<sup>a</sup> | 91 |

<sup>a</sup> Cross-sectional prevalence study.<sup>b</sup> Cohort study.<sup>c</sup> Case-control study.

### Assessment of the Motor development in children

Regarding the evaluation of motor development, we included five studies that had a control group to compare the scores in relation to the sample of premature / low weight individuals. The studies were grouped according to the tests used and an average of the standardized difference was calculated is shown in Fig. 2. The randomized effect model indicated the standardized mean difference of [-1.15 (95% CI -1.56, -0.73), I^2^ 80%]. Thus, it is evidenced an inferior motor development of children with LBW when compared to the control population. The results of motor development described by cross-sectional studies without a control group, were only described according to Table 2, as it was not possible to calculate the scores due to the high heterogeneity between the studies.

**Fig. 2.**
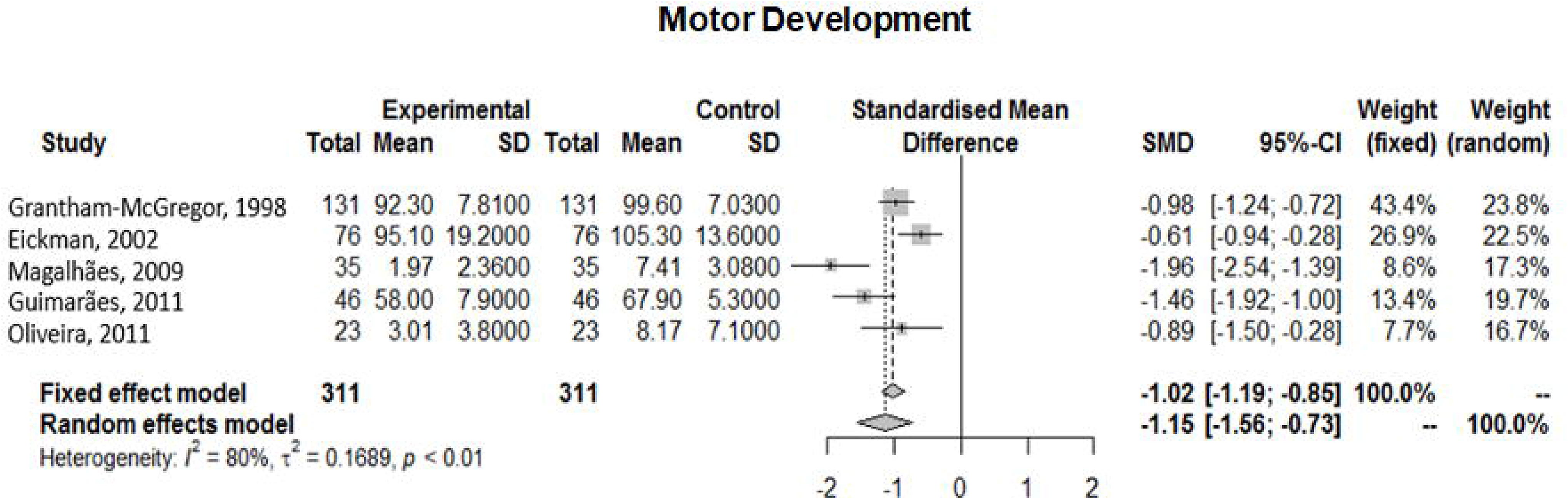
Forest plots showing motor development in children.

### Assessment of the cognitive development in children

To compare the results on cognitive development between the sample and the control population, five studies were included, according to the randomized effect model indicated the standardized mean difference of [-0,71 (95% CI -0.99, -0.44) I^2^ 67%], is shown in Fig. 3. Thusly, the studies indicated that children with LBW have lower cognitive development than term children. The results of cognitive development described by cross-sectional studies without a control group, were only described according to Table 2, as it was not possible to calculate the scores due to the high heterogeneity between the studies.

**Fig. 3.**
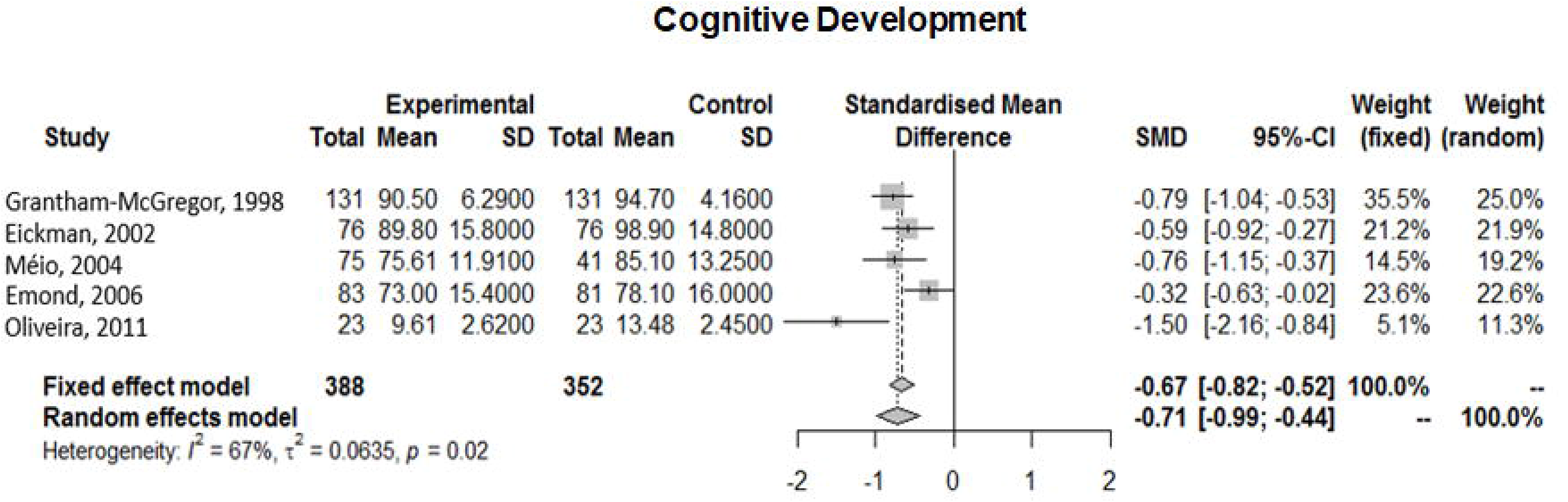
Forest plots showing cognitive development in children.

## Discussion

Our review reinforces that impaired motor and cognitive outcome is a significant long-term complication associated with low birth weight or malnutrition. The risk of impairment increases the lower the weight.

Sacchi et al.^6^ examined cognitive outcomes of preterm and term-born children who had intrauterine growth restriction (IUGR) and were small for gestational age (SGA) compared with children who were appropriate for gestational age (AGA) during the first 12 years of life. Their main findings were that growth vulnerabilities assessed antenatally (IUGR) and at the time of birth (SGA) were significantly associated with lower childhood cognitive outcomes in preterm and term-born children compared with children with AGA. These findings demonstrate the need for early interventions in these high risk groups as well as it reiterates the results of our study.

In a similar study, Kesavan et al.^7^ reinforces that IUGR infants suffer significant morbidity with immediate and long-term health consequences. These infants are at high risk throughout their life and should be carefully monitored at different stages to ensure timely interventions toward prevention and management of various disorders affecting most organ systems. Enhancing these data, Eickman et al.^23^ showed that LBW infants had poorer motor and cognitive development than ABW term infants, concluding that birth weight is considered a risk factor for outcome. These results enhance our analysis, where motor and cognitive delays were found in children with LBW.

In the same path, Courchia et al.^46^ reported cognitive outcomes of 632 preterm infants evaluated in a single center between 1980 and 2015. Significant cognitive impairment for all infants decreased by 9.4% (p = 0.015) across the study period. For larger infants (birthweight ≥ 750 g), significant impairment decreased by 14.6% (p = 0.002). In smaller infants (birthweight < 750 g) no significant changes were observed in cognitive outcomes over the study period. This cohort showed significant improvement in long term outcomes of infants since 1980.

Reinforcing the previously cited findings, Albu et al.^8^ and Sharma et al.^9^, associated IUGR with increased fetal and neonatal mortality and morbidity, being linked to immediate perinatal adverse events (prematurity, cerebral palsy, intrauterine fetal death, neonatal death). A selected study in our meta-analysis, Fuentefria et al.^28^ evidenced that VLBW preterm infants at 8 and 18 months corrected age showed significant differences in the neurodevelopment and growth pattern when compared with their full-term peers.

By exploring differences in neonatal body composition in a multi-ethnic population, and with the hypothesis that neonates from low and middle income countries (LAMIC) tend to have lower birth weight, Sletner et al^47^ found out that anthropometric measurements, such as abdominal circumference and ponderal index, were smaller in neonates from LAMIC origin when compared to western european neonates.

In a recent analysis of a Brazilian cohort.^43^ a comparison between 83 infants with birth weight ≤ 1,500 g, and 52 control infants was made. VLBW infants at 8 and 18 months corrected age showed significant differences in the neurodevelopment and growth pattern when compared with their full-term peers.

In 2013, a cross-sectional study included children aged 6-7 from a historical birth cohort with low birth weight (<2,500 g) infants evaluated cognitive and behavioral development of preterm and low birth weight newborns living in a disadvantageous socioeconomic environment at school age.^41^ The total intelligence quotient varied from 70 to 140. The borderline intelligence quotient was observed in 9.3% of the children and the Child Behavior Checklist indicated a predominance of social competence problems (27.8%, CI 19.2 to 37.9) compared with behavioral problems (15.5%, CI 8.9 to 24.2). Thus, their conclusion suggests that these infants are at risk for developing disturbances in early school age, such as mild cognitive deficits and behavioral disorders. This risk might increase under unfavorable socioeconomic conditions.

However, as limitations of our study, it is extremely important to point out that the majority of Brazilian follow-up studies are old and seem to demonstrate a lack of homogeneity. Although 24 Brazilian studies were selected, only five demonstrated homogeneity to perform meta-analysis. Yet, data when compared with the most recent world literature, mostly reiterate the relationship of prematurity with delayed neurological outcome, regardless of the scales used for analysis.^48,49^

With this review it was shown that impaired neurodevelopmental outcome is a significant long-term complication associated with low birth weight and malnutrition. Also, IUGR and SGA increases the risk of neurodevelopmental impairment. Thus, low birth weight represents risks to cognitive and motor development of children born, especially in the early years of life. From the perspective of public health, it is essential that pediatricians are aware of the neuropsychomotor development of these children for proper treatment and follow-up.

## Ethical Publication Statement

We confirm that we have read the Journal position on issues involved in ethical publication and affirm that this report is consistent with those guidelines.

## Data Availability

not applicable

## Funding

This research did not receive any other specific grant from funding agencies in the public, commercial, or not-for-profit sectors.

## Conflicts of interest

The authors declare no conflicts of interest.

## Notes

### Competing Interest Statement

The authors have declared no competing interest.

### Funding Statement

none

### Author Declarations

The article to be registered is a systematic review of the literature and does not require the signature of IRB.

